# Longitudinal Tracking and Construct Validity of a Single-Item Physical Activity Measure in the Women’s Healthy Ageing Project

**DOI:** 10.64898/2026.08.27.26361567

**Authors:** Daniel Corcoran, Cassandra Szoerke, Vasso Apostolopoulos, Jack Feehan

**Author notes:** Correspondence to. 225–245 Plenty Rd, Bundoora, Victoria, Australia 3083.

## Abstract

This study aimed to quantify the longitudinal tracking and cross-sectional construct validity of a single-item questionnaire measuring recreational physical activity frequency (RPAF) in the Women’s Healthy Ageing Project. At baseline, 474 participants aged 45-55 reported RPAF from 1993 to 2014. Longitudinal tracking of the RPAF item was assessed as a consecutive-wave and baseline-referenced measure using linear weighted kappa (LWK), Spearman correlations, exact agreement and within-one-category agreement. Construct validity in the form of convergent and known-group validity was assessed using the International Physical Activity Questionnaire (IPAQ) leisure activity domains, Short Form 36 physical function (SF-36-PF) subscale, Timed Up and Go (TUG), hand grip strength (HGS) and waist-to-height ratio (WhTR). 474 participants provided baseline RPAF data. Pairwise longitudinal samples ranged from 176 to 459 across the study. Consecutive-wave LWK ranged from 0.38 to 0.49, and Spearman correlations ranged from 0.44 to 0.57. Exact and within-category agreement ranged from 41.4%-50.8% and 72.0%-79.0%. Baseline-referenced LWK ranged from 0.22 to 0.47, with Spearman correlations of 0.29 to 0.56. RPAF correlated with total IPAQ leisure score (rs = 0.60), IPAQ walking score (rs = 0.58), SF-36-PF (rs = 0.33) and TUG score (rs = -0.25). No significant correlation was identified between RPAF, HGS or WhTR. RPAF discriminated known groups for WHO guideline-sufficient activity, SF-36-PF, and TUG fall risk. The RPAF item demonstrated fair-to-moderate agreement in consecutive waves, with weaker baseline-referenced tracking. Cross-sectional validity was highest with total IPAQ leisure activity. The item may provide a pragmatic measure for RPAF in women’s cohort studies.

## Introduction

Physical activity (PA) is an important determinant of healthy ageing, physical function and disease prevention. Current international PA guidelines recommend that older adults engage in regular PA that includes aerobic, balance, and strength activities to improve functional capacity (1). Longitudinal cohort research has previously reported positive findings for short-term objective PA measures (2); however, recent evidence on long-term objective PA assessment demonstrates limited adherence and missing data constraints (3), as well as technical limitations associated with measurement devices (4). As a result of these limitations, self-reported physical activity questionnaires (PAQ) are commonly utilised in clinical and public health research.

Brief or single-item PAQs offer a pragmatic alternative for PA measurement when objective measurement is not feasible. To date, several PAQs of varying design have been used to measure PA characteristics, including type, duration, intensity and volume (5); however, there remains no criterion standard PAQ that is universally agreed upon or implemented in health research (6). Previous longitudinal uses of PAQs include those in the Framingham Heart Study (7) and the Tromsø Study (8), whose data collection has spanned one or more decades. Fewer studies have investigated longitudinal PA measurement exclusively in women, with cohorts such as the Study of Women’s Health Across the Nation (9) and the Australian Longitudinal Study on Women’s Health (10) as prominent exceptions. Studies of PAQs used within these cohorts report varying short-term reliability and validity (11, 12) and moderate long-term reliability (13). Reliability within longitudinal cohorts, particularly those spanning decades, presents further challenges, as typical test-retest reliability assumes participant stability between measurements (14) during a time when PA behaviour is likely to change due to life transitions, particularly in women (15).

The Women’s Healthy Ageing Project (WHAP) is a multi-decade cohort that provides an opportunity to assess the longitudinal tracking and validity of a single-item measure of recreational physical activity frequency (RPAF) as women transition from mid-to late life. Earlier research using the WHAP RPAF item has examined PA change and short-term instrument properties in this cohort (16), but due to limited comparator outcomes in the early stage of the project, such as the International Physical Activity Questionnaire (IPAQ) (17), the long-term tracking and validity of the measure have yet to be established. In women, the transition period through midlife is particularly relevant, as women tend to experience greater longevity but higher rates of disability than their male counterparts (18). As midlife PA has been reported to improve late-life health (19), the measurement properties of a tool used to assess PA should be established prior to its implementation.

As objective measures are difficult to implement consistently across long-term cohorts (20), the suitability of subjective PAQs commonly used in large-cohort research remains unclear. This study aimed to assess the longitudinal tracking and cross-sectional validity of the subjective RPAF item in the WHAP. The findings may help determine whether a single-item PAQ is useful for assessing RPAF in longitudinal epidemiological research, particularly among women as they transition from midlife into later life.

## Methods

### Participants

Women included in this study were part of the Women’s Healthy Ageing Project (WHAP), an Australian longitudinal cohort study investigating a range of measures of women’s health across mid- and late-life (21). At the commencement of the study, women were aged 45-55 years and were followed up annually from 1993 to 1999, and then intermittently in 2002, 2004, 2012, and 2014. Participants were excluded from the study if they had undergone a hysterectomy prior to baseline intake. An extensive description of the participants, protocols and outcome measures collected during the WHAP has been detailed elsewhere (21). Ethical approval for the data collected during this study was granted by the University of Melbourne’s human research ethics committee (HREC) under the following identifiers: 931149X (1992–1999); 010528 and 010411 (2002–2009); 1034765 and 1339373 (2012–2016), and participants were briefed on the purpose and intent of the research prior to commencement. All participants gave voluntary informed consent prior to enrolment in the study.

### Self-reported recreational physical activity

In all years of the study, recreational PA frequency (RPAF) was measured using a single-item measure with seven ordinal frequency categories. Participants were asked “How often, if at all, do you participate in physical activities or sports for fitness or recreational purposes?” and responded on the following scale: Never, less than once a month, a few times a month, once a week, 2-3 times per week, 4-6 times per week and daily. Responses were collapsed into a five-category ordinal scale in line with previous validation research on the same cohort (16) and to improve the interpretability of the scale (22). Categories in the collapsed scale were: Less than once per week (0), once per week (1), 2-3 times per week (2), 4-6 times per week (3) and daily (4). In the later years of the study, participants also completed the IPAQ long form questionnaire (17). For the purpose of this study, only the scores relating to the leisure domains (items 20-26) of the IPAQ were used. In alignment with data processing recommendations, responses that exceeded 180 minutes per day were truncated to a maximum of 180 minutes (23). To calculate weekly Metabolic Equivalent of Task minutes (MET-minutes), the reported number of days performing each activity category (walking, moderate, and vigorous) was multiplied by the reported minutes per day using the following multipliers: 3.3 for walking, 4.0 for moderate and 8.0 for vigorous activity (17). While IPAQ was originally designed for adult populations, previous research has shown acceptable validity in older adults (24).

### Comparator measures and convergent validity

Convergent validity is a type of construct validity that assesses whether a specific instrument score is associated with other related construct measures (25). Functional and anthropometric outcome measures, including Timed Up and Go (TUG), handgrip strength (HGS), and waist-to-height ratio (WHtR), were analysed for their relationship with RPAF to assess convergent validity. TUG and HGS were included as measures of function for lower limb mobility (26) and muscle strength (27), respectively. Dominant hand HGS was recorded three times with a hand-held dynamometer, after which an average was calculated. WHtR was included as an anthropometric measurement in this analysis due to its relationship to disease risk and its preference over body mass index (BMI) in clinical screening (28). A subjective measure of physical function was also included in the form of the 36-Item Short Form Survey physical function (SF-36-PF) subscale (29). All comparator variables were modelled as continuous variables and were further transformed to create known-group comparator variables commonly used in healthcare contexts. Known-group validity is a type of construct validity that assesses whether an instrument can discriminate between distinct groups (30). A TUG threshold indicative of increased falls risk (≥12.5 seconds) (31) was used, along with a HGS threshold of <16 kg as an indicator of probable sarcopenia (27). WHtR scores were categorised according to usual (< 0.5) and increased (≥ 0.5) health risk associated with central adiposity (32). SF-36 PF was grouped into low (< 75) and high (≥ 75) function, based on previous studies that validated its use as a disability measure (33). Additionally, to determine whether the scale differentiated groups meeting World Health Organisation (WHO) sufficient activity recommendations, MET-minutes per week were categorised into under or over 600 MET-minutes (34). All validity measures were taken from the 2014 WHAP collection wave.

### Statistical analysis

Consecutive-wave (ten consecutive pairs) and baseline-referenced longitudinal tracking of the RPAF item was quantified using pairwise kappa calculations, Spearman’s rank order correlation coefficient (rs), exact and within-one-category agreements. As the RPAF scale was ordinal, a linear weighted kappa (LWK) was used to account for ordered responses and to penalise larger disagreements (35). Convergent validity between the RPAF item and comparator variables was assessed through Spearman’s rank correlation coefficient, and to compare clinically meaningful known groups, the Mann-Whitney U test was used. Values indicating agreement for LWK were 0-0.20, 0.21-0.40, 0.41-0.60, 0.61-0.80 and 0.81-1.0, indicating slight, fair, moderate, substantial, and almost perfect agreement (36). Values for Spearman’s rank correlation coefficient were interpreted as 0.00-0.10, 0.10-0.39, 0.40-0.69, 0.70-0.89 and 0.90-1.00, indicating negligible, weak, moderate, strong and very strong correlations, respectively (37). Non-parametric pointwise percentile bootstrap (1000 replications) confidence intervals were calculated for LWK, Spearman’s rank correlation coefficient and rank-biserial (rrb) statistics. As several outcomes were selected, the Benjamini-Hochberg procedure was applied across all validity assessments to control for false discovery rate (38). The probability value (p) was set at < 0.05 for all outcomes. As a sensitivity analysis, all tracking and validity measures were repeated using the original uncollapsed seven-category ordinal scale. All analyses were performed in R Statistical Software (v4.6.0) (39).

### Data availability

The WHAP cohort data used in this analysis are available through application to BioGrid Australia Limited (https://www.biogrid.org.au). BioGrid Australia and the data custodian approved permission for use.

## Results

Of the baseline 476 women in the cohort, 474 reported valid RPAF data in 1993, and by 2014 this number had reduced to 192. Consecutive-wave pairwise samples ranged from 176 to 459, and baseline-anchored samples ranged from 190 to 459. Relevant characteristics of the sample are detailed in Table 1. Full per-wave RPAF values are available in the Supplementary Material.

**Table 1.** 2014 cross-sectional cohort characteristics.

| **Variable** | **n** | **Mean (SD)** |
| --- | --- | --- |
| Age (years) | 192 | 71.98 (2.66) * |
| Recreational Physical Activity Frequency | 192 |  |
| Less than once per week | 45 (23.4%) |  |
| Once per week | 13 (6.8%) |  |
| 2-3 times per week | 48 (25.0%) |  |
| 4-6 times per week | 38 (19.8%) |  |
| Daily | 48 (25.0%) |  |
| IPAQ Leisure Total | 185 | 867.48 (1081.06) |
| Short-form 36 Physical Function | 189 | 71.53 (23.80) |
| Timed up-and-go (s) | 187 | 9.48 (2.50) |
| Hand grip strength (kg) | 187 | 22.12 (5.0) |
| Waist-to-height ratio | 188 | 0.54 (0.05) |

\*Age calculated from 2014 available RPAF reporting sample (n=192).

**Table 2.** Recreational physical activity frequency item consecutive-wave (ten pairs) tracking and agreement.

| **Wave** | **n** | **LWK (95% CI)** | **rs (95% CI)** | **Exact agreement %** | **Within-one category agreement %** |
| --- | --- | --- | --- | --- | --- |
| 1993-1994 | 459 | 0.47 (0.41, 0.53) | 0.56 (0.49, 0.63) | 49.89 | 74.51 |
| 1994-1995 | 450 | 0.45 (0.39, 0.51) | 0.53 (0.45, 0.61) | 46.67 | 76.22 |
| 1995-1996 | 447 | 0.47 (0.41, 0.52) | 0.55 (0.47, 0.63) | 48.77 | 75.84 |
| 1996-1997 | 443 | 0.43 (0.37, 0.49) | 0.51 (0.42, 0.58) | 47.86 | 72.46 |
| 1997-1998 | 436 | 0.42 (0.35, 0.48) | 0.49 (0.41, 0.57) | 45.64 | 72.02 |
| 1998-1999 | 431 | 0.48 (0.41, 0.54) | 0.54 (0.46, 0.62) | 50.81 | 75.64 |
| 1999-2002 | 259 | 0.40 (0.31, 0.48) | 0.48 (0.37, 0.59) | 44.40 | 74.52 |
| 2002-2004 | 229 | 0.38 (0.28, 0.46) | 0.44 (0.33, 0.56) | 42.36 | 77.29 |
| 2004-2012 | 181 | 0.38 (0.28, 0.47) | 0.47 (0.33, 0.60) | 41.44 | 74.59 |
| 2012-2014 | 176 | 0.49 (0.39, 0.58) | 0.57 (0.44, 0.68) | 48.86 | 78.98 |

**Table 3.** Recreational physical activity frequency item baseline-referenced tracking and agreement.

| **Years from baseline (1993)** | **n** | **LWK (95% CI)** | **rs (95% CI)** | **Exact agreement %** | **Within-one category agreement %** |
| --- | --- | --- | --- | --- | --- |
| 1 | 459 | 0.47 (0.41, 0.53) | 0.56 (0.49, 0.63) | 49.89 | 74.51 |
| 2 | 453 | 0.43 (0.37, 0.49) | 0.53 (0.45, 0.61) | 45.03 | 73.95 |
| 3 | 447 | 0.39 (0.32, 0.45) | 0.47 (0.39, 0.55) | 41.83 | 72.26 |
| 4 | 443 | 0.32 (0.25, 0.39) | 0.40 (0.31, 0.49) | 37.70 | 67.49 |
| 5 | 435 | 0.33 (0.27, 0.40) | 0.43 (0.35, 0.51) | 37.47 | 66.21 |
| 6 | 432 | 0.33 (0.26, 0.39) | 0.41 (0.32, 0.50) | 39.12 | 67.36 |
| 9 | 259 | 0.32 (0.23, 0.40) | 0.44 (0.33, 0.55) | 35.91 | 68.73 |
| 11 | 289 | 0.22 (0.14, 0.29) | 0.31 (0.21, 0.41) | 29.76 | 63.32 |
| 19 | 228 | 0.22 (0.14, 0.31) | 0.31 (0.18, 0.42) | 30.70 | 61.40 |
| 21 | 190 | 0.23 (0.13, 0.33) | 0.29 (0.16, 0.43) | 33.68 | 60.53 |

**Table 4.** Recreational physical activity frequency item convergent validity.

| **Variables** | **n** | **rs (95% CI)** | **p\*** |
| --- | --- | --- | --- |
| IPAQ Leisure Total MET-minutes | 185 | 0.60 (0.49, 0.69) | < 0.01 |
| IPAQ Walking MET-minutes | 185 | 0.58 (0.47, 0.68) | < 0.01 |
| IPAQ Moderate MET-minutes | 185 | 0.21 (0.10, 0.33) | < 0.01 |
| IPAQ Vigorous MET-minutes | 185 | 0.25 (0.13, 0.36) | < 0.01 |
| Timed-up-and-go | 187 | -0.25 (-0.38, -0.10) | < 0.01 |
| Hand grip strength | 187 | 0.12 (-0.03, 0.25) | 0.15 |
| Short-form 36 Physical Function | 189 | 0.33 (0.18, 0.46) | < 0.01 |
| Waist-to-height ratio | 188 | -0.07 (-0.22, 0.09) | 0.38 |

\*Benjamini-Hochberg adjusted p-value

**Table 5.** Recreational physical activity frequency item known-groups validity.

| **Known group** | **n**<br>**(Group 1/2)** | **MW U** | **Median (IQR) per**<br>**group** | **rrb** | **p\*** |
| --- | --- | --- | --- | --- | --- |
| WHO sufficient activity<br>(MET-minutes < 600, ≥ 600) | 185<br>(110/75) | 1809.5 | 1 (0-3)/3 (2-4) | 0.56 | < 0.01 |
| Short-form 36 Physical Function<br>(< 75, ≥ 75) | 189<br>(77/112) | 2891.5 | 2 (0-3)/3 (2-4) | 0.33 | < 0.01 |
| Timed-up-and-go<br>(< 12.5s, ≥ 12.5s) | 187<br>(177/10) | 1338.0 | 2 (1-4)/0 (0-1.5) | -0.51 | 0.01 |
| Hand grip strength<br>(< 16kg, ≥ 16kg) | 187<br>(16/171) | 1431.5 | 2 (1.75-3.25)/2 (1-3.5) | -0.05 | 0.75 |
| WHtR risk category<br>(< 0.5, ≥ 0.5) | 188<br>(35/153) | 2561.0 | 2 (0.5-3)/2 (1-3) | 0.04 | 0.74 |

Key: Positive rank-biserial correlation indicates higher RPAF in Group 2 than Group 1; negative values indicate lower RPAF in Group 2. \*Benjamini-Hochberg adjusted p-value.

### Consecutive-wave longitudinal tracking

Pairwise LWK estimates ranged from 0.38 to 0.49 across all waves, with the highest (0.49) and lowest (0.38) values occurring in 2012-2014 and 2002-2004, respectively, indicating fair to moderate agreement. Mean LWK estimates were higher when waves were annual (0.45) and decreased stepwise as years between data collection increased (0.43, 0.40, 0.38 for 2, 3, and 8-year intervals, respectively). Spearman’s rank correlation coefficient ranged from 0.44 to 0.57, highest (0.57) and lowest (0.44), matching the same pattern as LWK estimates. Exact agreement ranged from 41.44% to 50.81% across all waves, and within-one-category agreement ranged from 72.02% to 78.98%, indicating that while exact agreement was low, it was consistent (> 70%) within one ordinal level.

### Baseline-referenced longitudinal tracking

LWK estimates ranged from 0.22 to 0.47 across all years from baseline. LWK estimates declined considerably (0.32 to 0.22) beyond 11 years from baseline. Spearman’s rank correlation coefficient showed a similar pattern, with the lowest estimate (rs = 0.29) at 21 years from baseline. Exact agreement was lowest (29.76%) and highest (49.89%) at 11 and 1 year from baseline, respectively. Within-one-category agreement followed similar patterns to LWK and Spearman’s rank correlation, with the lowest value (60.53%) occurring at 21 years from baseline. All estimates declined as years from baseline increased, although the decline was not monotonic.

### Cross-sectional validity

RPAF was significantly correlated with IPAQ leisure domain MET-minutes (rs = 0.21-0.60). The strongest correlation occurred with IPAQ Leisure Total MET-minutes (rs = 0.60) and was followed by IPAQ Walking MET-minutes (rs = 0.58). Additional significant, although weaker, correlations were identified with SF-36-PF (rs = 0.33) and TUG (rs = - 0.25). No significant correlation was present between RPAF and HGS or WHtR.

Compared with known groups, RPAF showed significant differences in WHO guideline-sufficient activity, SF-36 PF high and low function, and TUG-related falls risk.Participants reporting higher RPAF scores had higher SF-36 PF scores (U = 2891.5, rrb = 0.33, 95% CI 0.18, 0.47) and lower TUG times (U = 1338.0, rrb = -0.51, 95% CI -0.76, - 0.18). Significant differences were also identified in meeting WHO sufficient activity thresholds (U = 1809.5, rrb = 0.56, 95% CI 0.44, 0.69). No significant differences were identified in HGS or WHtR groups.

### Sensitivity Analysis

Results from the sensitivity analysis using the original uncollapsed seven-category ordinal scale were predominantly unchanged, with the largest variations noted in exact and within-one-category analyses. Full details are available in the Supplementary Material (Tables S2-S6).

## Discussion

Previous studies have explored the reliability of various PAQs (5, 15); however, the results from this study should not be interpreted as standard ‘reliability’ performed in psychometric testing, as those tests are typically impacted by administration interval (40) and likely do not factor in behavioural change occurring as women transition from mid-to-late life. The results from the RPAF item used in the WHAP are better considered as longitudinal behavioural (15) or trait-tracking (41) rather than test-retest reliability. The findings from this study support traditional statistics used to assess longitudinal tracking and agreement (i.e., rs and kappa statistics) but additionally report exact and within-one-category agreement. The key findings support using the RPAF measurement item as a brief PAQ to assess behavioural or habitual RPAF among mid- to late-life women when administered repeatedly. Supporting previous PAQ findings (5), the WHAP RPAF item demonstrated fair to moderate longitudinal agreement and tracking (LWK 0.38 – 0.49, rs = 0.44 – 0.57), and while exact agreement was poor (41.4% - 50.8%), changes were consistently observed within a single category (> 70% at all intervals), indicating that paired responses were either identical or within one category. Findings for baseline-referenced tracking were considerably weaker, with all estimates declining as years from baseline (1993) increased.

Due to methodological differences and the lack of studies reporting both reliability and validity (5), direct comparisons between PAQs are difficult. Among studies using similar statistical methods, outcomes are consistent, though reported in magnitudes differing from those found with the current RPAF item. Gill et al. (2012) (42) used absolute and relative PA items to assess reliability over a 1-week period. They found that an absolute (total) PA item demonstrated higher agreement (k = 0.75, 95% CI 0.60, 0.91) than a relative (peer-compared) item (k = 0.56, 95% CI 0.30, 0.82). Another PAQ cohort study also displayed fair-to-good agreement over a maximum five-month interval, noting that an hours per week of ‘sports’ item displayed higher agreement (k = 0.57, 95% CI 0.47, 0.68) than hours per week of ‘moderate’ activity item (k = 0.30, 95% CI 0.23, 0.37) (43). While short-term studies have related measurement properties, longer-term studies can reflect both measurement and behavioural variability (15). In the WHAP, baseline-referenced LWK declined across a 21-year period from 0.47 to 0.23. Other long-term studies have reported similar kappa estimates, such as those from Morseth et al. (2011), who reported estimates of 0.41 and 0.29 for 7- and 28-year intervals,respectively; and Aggio et al. (2017) (44), who reported estimates of 0.26, 0.23 and 0.24 for 12, 16 and 20-year intervals, respectively. The contrasting findings between consecutive-wave and baseline-referenced outcomes in this analysis suggest that the WHAP RPAF item is potentially useful if administered repeatedly but, in agreement with existing research, displays poorer longitudinal tracking potential as the years between administrations increase.

Much like longitudinal tracking, validity assessments of PAQs also show limited methodological consistency, with studies often validating against physical function outcomes, device-based measures or other PAQs (5). Gill et al. (2012) (42) reported convergent validity across a range of physical function measures, with negligible to weak (rs = 0.10-0.33) and weak to moderate (rs = 0.28-0.57) associations for absolute and relative PA items, respectively, with highest validity for walking and gait path.Similar correlations have been identified with physical function (rs = 0.40), mobility frequency (rs = 0.59) and walking difficulty (rs = -0.61) (45). A recent study validating a single-item lifestyle PA item used the IPAQ in a similar manner to this study (46). The study reported that IPAQ moderate PA (rs = 0.27) and SF-36 PF (rs = 0.36) showed correlations very similar to those of the RPAF item used in the WHAP, and additional similarities were observed in IPAQ Walking (rs = 0.26) and IPAQ Vigorous PA (rs = 0.39), though at lower magnitudes (46). While convergent validity is relatively common, known-group validity is less reported. Existing evidence on known-group validity has demonstrated that higher lifestyle activity differentiated between IPAQ intensity domains, SF-36 subscales, BMI and various functional measures (46, 47). In functional findings comparable to this study, lifestyle activity level differentiated fall risk as measured by chair stand, with higher activity associated with improved performance (47).

### Research implications

The results in this paper build upon existing evidence for single-item or brief PAQs, including those suggesting that PAQs capable of ranking activity groups are likely suitable for longitudinal association analysis (48). To date, very few studies have collected PA data across two decades at the frequency of the WHAP, and fewer have focused on the mid-to-late life transition in women. The findings in this study support the use of the RPAF item as a tracking measure of habitual recreational PA across 1- to 8-year collection intervals, with validity relative to lower-limb function tests, subjective physical function, and commonly used subjective physical activity questionnaires in line with previous existing research (46, 47). Future research exploring single-item or brief PAQs should validate subjective PA against objective energy expenditure measures to improve their interpretative value.

### Limitations

While the RPAF item is useful for ranking purposes based on the analysis in this study, several limitations are present. The RPAF measure cannot distinguish between true behavioural change and measurement variability, and it considers frequency only; it should not be viewed as a measure of intensity, energy expenditure, or adherence to PA guidelines. Additionally, primary correlations were with other IPAQ leisure items that are not objective measures and show weak correlations with such measures (17). Further, some known group sizes were small, which reduces the precision of point estimates. As with most cohort research, the findings from this study may have limited generalisability external to the WHAP sample. Additionally, later-wave participation in the study may represent selective survival or retention that may exclude participants who differ in health status or PA behaviour stability compared to those who were retained.

## Conclusion

The item used within the WHAP provides a brief self-reported ordinal measure of RPAF frequency among mid-to-late life women. The measure demonstrated fair-to-moderate agreement and tracking with repeated administration, along with cross-sectional convergent validity with IPAQ leisure and walking items and SF-36 subjective physical function. The item also showed cross-sectional known-group validity for WHO guideline-sufficient activity, SF-36 PF, and TUG-related fall risk. The findings in this study support the use of the item as a brief longitudinal PA tracking measure, likely useful for research incorporating repeated administrations. Future investigations should focus on the relationship of similar items to existing objective behavioural or habitual PA measures.

## Data Availability

The cohort data used in this analysis are available through application to BioGrid Australia Limited (https://www.biogrid.org.au).

