## Supplementary Material for "Longitudinal Tracking and Construct Validity of a Single-Item Physical Activity Measure in the Women’s Healthy Ageing Project"

*Supplementary Table S1. Recreational Physical Activity Frequency Item Per Wave Distribution*

| Year | n (% of 1993) | Missing RPAF response | Less than once/week (%) | Once/week (%) | 2–3 times/week (%) | 4–6 times/week (%) | Daily (%) |
|---|---|---|---|---|---|---|---|
| **1993** | 474 (99.6%) | 2 | 142 (30.0%) | 51 (10.8%) | 119 (25.1%) | 71 (15.0%) | 91 (19.2%) |
| **1994** | 460 (96.66%) | 16 | 127 (27.6%) | 48 (10.4%) | 122 (26.5%) | 73 (15.9%) | 90 (19.6%) |
| **1995** | 454 (95.4%) | 22 | 117 (25.8%) | 51 (11.2%) | 122 (26.9%) | 75 (16.5%) | 89 (19.6%) |
| **1996** | 448 (94.1%) | 28 | 112 (25.0%) | 45 (10.0%) | 104 (23.2%) | 88 (19.6%) | 99 (22.1%) |
| **1997** | 444 (93.3%) | 32 | 112 (25.2%) | 37 (8.3%) | 109 (24.5%) | 92 (20.7%) | 94 (21.2%) |
| **1998** | 436 (91.6%) | 40 | 115 (26.4%) | 39 (8.9%) | 99 (22.7%) | 77 (17.7%) | 106 (24.3%) |
| **1999** | 433 (91.0%) | 43 | 100 (23.1%) | 38 (8.8%) | 108 (24.9%) | 81 (18.7%) | 106 (24.5%) |
| **2002** | 260 (54.6%) | 216 | 46 (17.7%) | 24 (9.2%) | 78 (30.0%) | 58 (22.3%) | 54 (20.8%) |
| **2004** | 290 (60.9%) | 186 | 52 (17.9%) | 23 (7.9%) | 87 (30.0%) | 75 (25.9%) | 53 (18.3%) |
| **2012** | 229 (48.1% | 247 | 59 (25.8%) | 20 (8.7%) | 52 (22.7%) | 54 (23.6%) | 44 (19.2%) |
| **2014** | 192 (40.3%) | 284 | 45 (23.4%) | 13 (6.8%) | 48 (25.0%) | 38 (19.8%) | 48 (25.0%) |

*Supplementary Table S2. Seven-Category (uncollapsed) Recreational Physical Activity Frequency Item consecutive-wave (ten pairs) tracking and agreement.*

| Wave | n | LWK (95% CI) | rs (95% CI) | Exact agreement % | Within-one category agreement % |
|---|---|---|---|---|---|
| 1993-1994 | 459 | 0.46 (0.39, 0.51) | 0.56 (0.48, 0.63) | 43.79 | 67.10 |
| 1994-1995 | 450 | 0.46 (0.40, 0.53) | 0.54 (0.46, 0.61) | 43.11 | 71.56 |
| 1995-1996 | 447 | 0.46 (0.40, 0.54) | 0.55 (0.46, 0.63) | 44.74 | 69.57 |
| 1996-1997 | 443 | 0.43 (0.36, 0.49) | 0.51 (0.42, 0.58) | 44.02 | 67.27 |
| 1997-1998 | 436 | 0.41 (0.34, 0.48) | 0.50 (0.41, 0.58) | 42.66 | 66.51 |
| 1998-1999 | 431 | 0.49 (0.42, 0.55) | 0.55 (0.47, 0.62) | 47.56 | 71.00 |
| 1999-2002 | 259 | 0.41 (0.31, 0.49) | 0.49 (0.38, 0.59) | 41.70 | 70.27 |
| 2002-2004 | 229 | 0.37 (0.27, 0.45) | 0.44 (0.33, 0.56) | 37.99 | 72.05 |
| 2004-2012 | 181 | 0.36 (0.26, 0.46) | 0.47 (0.34, 0.59) | 37.57 | 69.06 |
| 2012-2014 | 176 | 0.44 (0.35, 0.54) | 0.57 (0.44, 0.69) | 44.89 | 70.45 |

*Supplementary Table S3. Seven-Category (uncollapsed) Recreational physical activity frequency item baseline-referenced tracking and agreement.*

| Years from baseline (1993) | n | LWK (95% CI) | rs (95% CI) | Exact agreement % | Within-one category agreement % |
|---|---|---|---|---|---|
| 1 | 459 | 0.46 (0.39, 0.51) | 0.56 (0.48, 0.63) | 43.79 | 67.10 |
| 2 | 453 | 0.43 (0.36, 0.49) | 0.54 (0.46, 0.61) | 39.96 | 66.00 |
| 3 | 447 | 0.40 (0.34, 0.47) | 0.48 (0.40, 0.56) | 37.81 | 64.21 |
| 4 | 443 | 0.32 (0.25, 0.39) | 0.41 (0.32, 0.49) | 33.63 | 60.27 |
| 5 | 435 | 0.34 (0.28, 0.41) | 0.44 (0.36, 0.51) | 33.56 | 58.85 |
| 6 | 432 | 0.34 (0.27, 0.40) | 0.41 (0.33, 0.50) | 34.95 | 60.19 |
| 9 | 259 | 0.35 (0.27, 0.43) | 0.45 (0.35, 0.56) | 32.43 | 61.39 |
| 11 | 289 | 0.22 (0.14, 0.29) | 0.31 (0.21, 0.41) | 23.18 | 56.06 |
| 19 | 228 | 0.22 (0.13, 0.31) | 0.31 (0.19, 0.43) | 26.75 | 51.32 |
| 21 | 190 | 0.21 (0.11, 0.31) | 0.29 (0.15, 0.42) | 30.00 | 53.68 |

*Supplementary Table S4. Seven-Category (uncollapsed) Recreational physical activity frequency item convergent validity*

| Variables | n | rs (95% CI) | p* |
|---|---|---|---|
| IPAQ Leisure Total MET-minutes | 185 | 0.60 (0.49, 0.69) | < 0.01 |
| IPAQ Walking MET-minutes | 185 | 0.59 (0.48, 0.68) | < 0.01 |
| IPAQ Moderate MET-minutes | 185 | 0.22 (0.10, 0.33) | < 0.01 |
| IPAQ Vigorous MET-minutes | 185 | 0.26 (0.14, 0.36) | < 0.01 |
| Timed-up-and-go | 187 | -0.25 (-0.39, -0.11) | < 0.01 |
| Hand grip strength | 187 | 0.12 (-0.02, 0.26) | 0.13 |
| Short-form 36 Physical Function | 189 | 0.34 (0.20, 0.47) | < 0.01 |
| Waist-to-height ratio | 188 | -0.08 (-0.22, 0.08) | 0.34 |

*Benjamini-Hochberg adjusted p-value

*Supplementary Table S5. Seven-Category (uncollapsed) Recreational physical activity frequency item known-groups validity*

| Known group | n (Group 1/2) | MW U | Median (IQR) per group | rrb | p* |
|---|---|---|---|---|---|
| WHO sufficient activity (MET-minutes < 600, ≥ 600) | 185 (110/75) | 1815.0 | 3 (0-5)/5 (4-6) | 0.56 | < 0.01 |
| Short-form 36 Physical Function (< 75, ≥ 75) | 189 (77/112) | 2822.5 | 4 (0-5)/5 (4-6) | 0.35 | < 0.01 |
| Timed-up-and-go (< 12.5s, ≥ 12.5s) | 187 (177/10) | 1352.5 | 4 (3-6)/0 (0-3.5) | -0.53 | 0.01 |
| Hand grip strength (< 16kg, ≥ 16kg) | 187 (16/171) | 1421.5 | 4 (3.75-5.25)/4 (3-5.5) | -0.04 | 0.79 |
| WHtR risk category (< 0.5, ≥ 0.5) | 188 (35/153) | 2602.0 | 4 (2.5-5)/4 (3-5) | 0.03 | 0.79 |

Key: Positive rank-biserial correlation indicates higher RPAF in Group 2 than Group 1; negative values indicate lower RPAF in Group 2. *Benjamini-Hochberg adjusted p-value. =

*Supplementary Table S6. Comparison of five (collapsed) and seven (uncollapsed) category recreational physical activity frequency response scales.*

| Analysis | Estimate | Five-category response scale | Seven-category response scale |
|---|---|---|---|
| Consecutive-wave tracking and agreement | LWK range | 0.38–0.49 | 0.36–0.49 |
| | Spearman ρ range | 0.44–0.57 | 0.44–0.57 |
| | Exact agreement range | 41.4%–50.8% | 37.6%–47.6% |
| | Within-one-category range | 72.0%–79.0% | 66.5%–72.1% |
| Baseline-referenced tracking and agreement | LWK range | 0.22–0.47 | 0.21–0.46 |
| | Spearman ρ range | 0.29–0.56 | 0.29–0.56 |
| | Exact agreement range | 29.8%–49.9% | 23.2%–43.8% |
| | Within-one-category range | 60.5%–74.5% | 51.3%–67.1% |
| Convergent validity | BH-significant correlations | 6/8 | Identical 6/8 |
| Known-group validity | BH-significant comparisons | 3/5 | Identical 3/5 |
